# Developing and validating a multi-criteria decision analytic tool to assess the value of cancer clinical trials

**DOI:** 10.1101/2022.12.07.22283233

**Authors:** Piers Gillett, Robert K Mahar, Nancy R Tran, Mark Rosenthal, Maarten IJzerman

**Affiliations:** Cancer Health Services Research Unit, Centre for Health Policy, Melbourne School of Population and Global Health, University of Melbourne, Australia; Biostatistics Unit, Centre for Epidemiology and Biostatistics, Melbourne School of Population and Global Health, University of Melbourne, Australia; Sir Peter MacCallum Department of Medical Oncology, University of Melbourne, Australia; Department of Medical Oncology, The Royal Melbourne Hospital, Australia

**Keywords:** Multi-criteria decision analysis, clinical trials, cancer, decision tool, decision analysis, prioritisation, portfolio

## Abstract

**Background:** Demonstrating safety and efficacy of new medical treatments requires clinical trials. But clinical trials are costly and may not provide value proportionate to their costs. In health systems with limited resources, it is important to identify the trials with the highest value. Tools exist to assess elements of a clinical trial such as statistical validity but are not wholistic in their valuation of a clinical trial. This study aims to develop a measure of clinical trials value and provide an online tool for clinical trial prioritisation.

**Methods:** A search of the academic and grey literature and expert consultation was undertaken to identify a set of metrics to aid clinical trial valuation using multi-criteria decision analysis. Swing weighting and ranking exercises were used to calculate appropriate weights of each of the included metrics and to estimate the partial-value function for each underlying metric. The set of metrics and their respective weights were applied to the results of six different clinical trials to calculate their value.

**Results:** Seven metrics were identified: ‘unmet need’, ‘size of target population’, ‘eligible participants can access the trial’, ‘patient outcomes’, ‘total trial cost’, ‘academic impact’ and ‘use of trial results’. The survey had 80 complete sets of responses (51% response rate). A trial designed to address an ‘Unmet Need’ was most commonly ranked as the most important with a weight of 24.4%, followed by trials demonstrating improved ‘Patient Outcomes’ with a weight of 21.2%. The value calculated for each trial allowed for their clear delineation and thus a final value ranking for each of the six trials.

**Conclusion:** We confirmed that the use of the decision tool for valuing clinical trials is feasible and that the results are face valid based on the evaluation of six trials. A proof-of-concept applying this tool to a larger set of trials with an external validation is currently underway.

## 1 Introduction

Clinical trials play a key role in establishing safety and efficacy of new modes of medical care. Beyond this, trials also generate externalities that accrue to different stakeholders, each of whom may value a trial in ways that go beyond safety or efficacy. Low uncertainty around treatment efficacies benefits future patients and society in general, is a requirement of health economic models that justify or redirect resources to areas of need and is essential for market authorisation and policy making [1].

Clinical trials are often costly, yet it remains difficult to establish whether a trial produces value commensurate with their cost. The respective average cost of phase I, II, and III trials of investigational compounds globally reached approximately $25.3M, $58.6M and $255.4M in 2013 U.S. dollars [2]. In Australia, the cost of early-stage/phase I trials, approximately 28% lower than the U.S. [3], is also high. This leads to the presumption that, in a society with limited resources, only trials that pose the highest value for society should be prioritised [4]. The American Society of Clinical Oncology (ASCO) [5] and the European Society of Medical Oncology (ESMO) [6] each provide frameworks to quantify gains of new cancer treatments based on patient survival, treatment toxicity and quality of life as determined in a clinical trial. While both scales have value, the outputs of the ASCO and ESMO frameworks are not well correlated. Moreover, a negative correlation between ASCO medical benefit scores and monthly drug costs was reported [7]. Some have found that most drugs enter the market without evidence of survival gain [8]. With high trial costs and relatively few trials providing clinical results that meet the magnitude of clinical benefit, it is important to establish a value threshold inclusive of diverse stakeholder preferences and sufficient to change clinical practice in order to justify high costs of the trial in the first place.

Because trial results can be used to inform future research directions, inform policy, or prompt changes in clinical practice, trial conclusions must be scientifically robust. Efforts to address this have produced tools such as the Cochrane risk-of-bias tool [9], the Jadad score [10], a general tool assessing methodological quality of a trial [11], a tool for methodological strength of orthopaedic surgery-focused trials [12] and the Delphi List [13]. Yet these tools tend to focus on relatively narrow definitions or a specific element of clinical trial value and none provide a broad measure of clinical trial value incorporating all stakeholder views.

Multi-criteria decision analysis (MCDA) is a method that enables users to assess conflicting criteria together to assist in decision making [14]. Each MCDA criterion represents something that at least some stakeholders consider important in decision making. MCDA helps to jointly assess each criterion. MCDA is a general concept and can be used for a variety of decision problems, including supporting patient choices and portfolio management [15]–[17]. Many different approaches could be considered MCDA, although each approach consists of the same core method. This core method includes selecting decision alternatives (different trials in our study), identifying criteria relevant for the decision, gauging the performance of the decision alternatives with respect to the criteria, weighting the criteria in terms of their importance and aggregating the weighted criteria into a single value [18]. Some of these methods allow the decision maker to explicitly quantify the value of each decision based on each different criterion (19).

In this study we implemented MCDA using swing weighting and ranking methods where responses from a range of representative stakeholders relevant for the design, conduct and results of clinical trials were collected. Stakeholders we considered included clinicians, statisticians, scientists, regulators, clinical trial unit managers, and consumer representatives. We recorded their opinions on which trial characteristics (herein referred to as ‘metrics’) of clinical trials encapsulated trial value and the relative value of each metric. What follows is a summary of how we developed an MCDA-based decision tool to evaluate cancer clinical trials and the application of the tool to retrospectively evaluate a portfolio of cancer clinical trials.

## 2 Methods

### 2.1 Step 1: Identifying metrics

A search through academic and grey literature was undertaken to establish a ‘starting set’ of metrics. Two reports from RAND Europe, the first by Guthrie et al. [19] and the second by Deshpande et al. 2018 (20) were identified, from which were identified, from which an initial set of eight metrics were tentatively selected.

These initial eight metrics were then augmented after consulting relevant stakeholders including health economists, oncologists, statisticians and operations researchers. Interviewees were presented the aims of our study and asked to suggest metrics they thought would “best represent clinical trial value”. Responses were either recorded manually or using Poll Everywhere, a web-based polling service [21]. Stakeholders were then asked to rank how representative of trial value the augmented set of metrics were for each of clinical trial phases I, II and III respectively using Poll Everywhere.

The augmented starting set of metrics was then considered in the context of three real-world clinical trials. An expert committee comprising clinicians, researchers, governmental representatives and clinical trialists were presented the three real-world trials, each from a different phase and with different interventions and asked to nominate which they considered the most valuable. Attendees were given a 10-minute time limit and could ask questions throughout. Participants were then asked to identify the key metric that, should its value change significantly, would change their decision. This was then repeated for each of the eleven metrics of the augmented set.

Following the expert consultations, we conducted a series of face-to-face interviews with patient advocates and health regulators as representatives of users. These interviews were conducted as open discussions between the research team and participants. We presented the participants with a list of metrics established through the previous rounds of polling and interviews. Participants were asked to rank the metrics representing trial value from ‘most representative’ of value to ‘least representative’. Participants were asked to talk through their process of ranking, so that we could record qualitative information about the gaps between metrics and their reasoning for each metrics position. A final set of seven metrics was agreed upon by the co-authors and steering group that was judged to best balance the different interests of all stakeholder groups.

### 2.2 Step 2: MCDA weights elicitation survey

Using the final set of metrics, a survey was designed using Qualtrics [22] to collect relative numeric weights of the metrics from a range of stakeholders. Full details of the survey can be found in Online Resource 1. The first four questions were all multiple choice and asked for the respondent’s involvement in clinical trials, whether they have any paid affiliations with the pharmaceutical or biomedical industry, their level of experience designing or running a clinical trial and their country of residence. Participants were then provided with definitions of the seven metrics being assessed and the range of likely values these metrics could take. They were then asked to rank the metrics from most to least important using swing weighting. Swing weighting requires respondents to provide rankings based upon the value gained should a metric improve from its worst possible outcome to its best. Respondents were then asked to provide estimates of weights (0–99) of each metric relative to the top ranked metric which had a weight of 100. The survey was open between 1 May 2020 and 31 July 2020. Participants were identified through professional networks and discussions with interested stakeholders. Each invited participant was sent an email containing survey details and a link to the survey. Those receiving an email invitation were also invited to forward the survey link to anyone whom they believed might be interested in participating. Consent was requested before participants could complete the survey.

### 2.3 Step 3: Eliciting partial value functions

We used partial value functions (PVFs) to transform each respective metric to a common ‘value’ scale ranging from 0 to 100. For example, the ‘patient outcomes’ metric is measured in terms of months of survival, which is then transformed using the appropriate PVF onto the common ‘value’ scale. For each metric, we established the functional form of each PVF through a series of interviews using the bisection method. The bisection method required that for each metric, respondents were asked at what point within the range of possible values for a given metric, with its worst possible outcome corresponding to a value of 0 and its best possible outcome a value of 100, would be equivalent to a value of 50. Alternatively, at what measure of the metric, was an increase from its worst possible measure to that point, equal in value to an increase from that point to its best possible outcome. All other metrics were held constant throughout.

Interviews to elicit PVFs were conducted with six people, a molecular scientist, consumer representative, health economist, clinical trials nurse, oncologist and a clinical project manager. Each interview began with an explanation of the project and why PVFs were required. The bisection method was explained in detail. Participants were then provided with examples and given the opportunity to practice on a simple example. We then explained the definition of each metric and eliciting their PVFs in turn. As participants responded, results were shown graphically to provide the participants with constant feedback. Results were recorded manually throughout.

The PVF for each metric was calculated using the average midpoint of the possible values of a metric, that corresponded to a value of 50. The parameters of a linear function were then calculated to create a straight line from the lowest value point of the metric to the average midpoint. This was repeated from the midpoint to the point of greatest value for each metric. This created ‘bent-stick’ style PVF. The equations for each PVF can be seen in Online Resource 2.

### 2.4 Step 4: Data analysis

Of the completed surveys, responses were separated into one of two categories, concordant or discordant. Concordant responses are those where metric rankings matched the descending order of metric weights.

The metric weights provided in the concordant data were standardised so they all fit on the same scale, 0 to 100. The standardisation was carried out using **Error! Reference source not found**. where ‘*w*_*i*_’ is the weight provided by the respondent for a given metric.

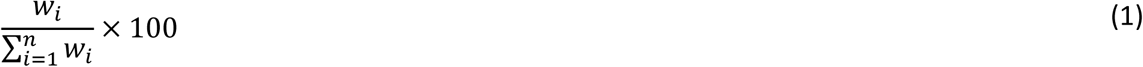

The weights provided by respondents in discordant responses were replaced with a set of weights calculated using the reciprocal of rank formula (Eq. 2) [23]. The formula is based on the number of metrics, *n* and the rank *k* given by the respondent for that metric (out of *n*). The sum of the standardised weights for each individual response equalled 100.

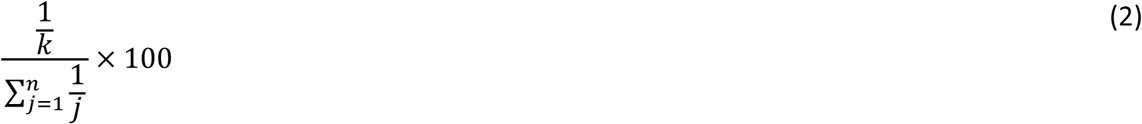

Data from the concordant and, formerly, discordant categories were then combined.

Using the standardised weights calculated for each metric, an average standardised weight was calculated for each metric using the combined discordant and concordant data. Analysis was carried out using R version 4.0.2 [24].

#### 2.4.1 Development of an online Shiny app

A publicly available Shiny web app was developed to facilitate the implementation and further development, of the MCDA tool using the results of our study. The Shiny app allows users to manually input or upload a CSV file containing data on the clinical trial metrics. The Shiny web app applies the PVFs and weightings to the data and aggregates the results to produce a single value measure for each trial. The web app can be viewed at Gillett et al., 2020 [25].

#### 2.4.2 Validation of the app

Data from six previously completed cancer clinical trials were used as a proof-of-concept of the decision tool. A value for each trial was calculated using the standardised weights and the PVF as programmed in the online tool. Data pertaining to our metrics of interest was collected and, where information for a specific metric was not available, it was filled in with a median value or a value from a similar trial as a substitute. Trials selected covered a range of interventions and are presented in Table 1. The aggregate trial value was calculated for each trial by summing the inputs for each metric, adjusted by the survey derived weights. The results were plotted and highlight the total trial value as well as the respective contributions of each metric.

**Table 1.**
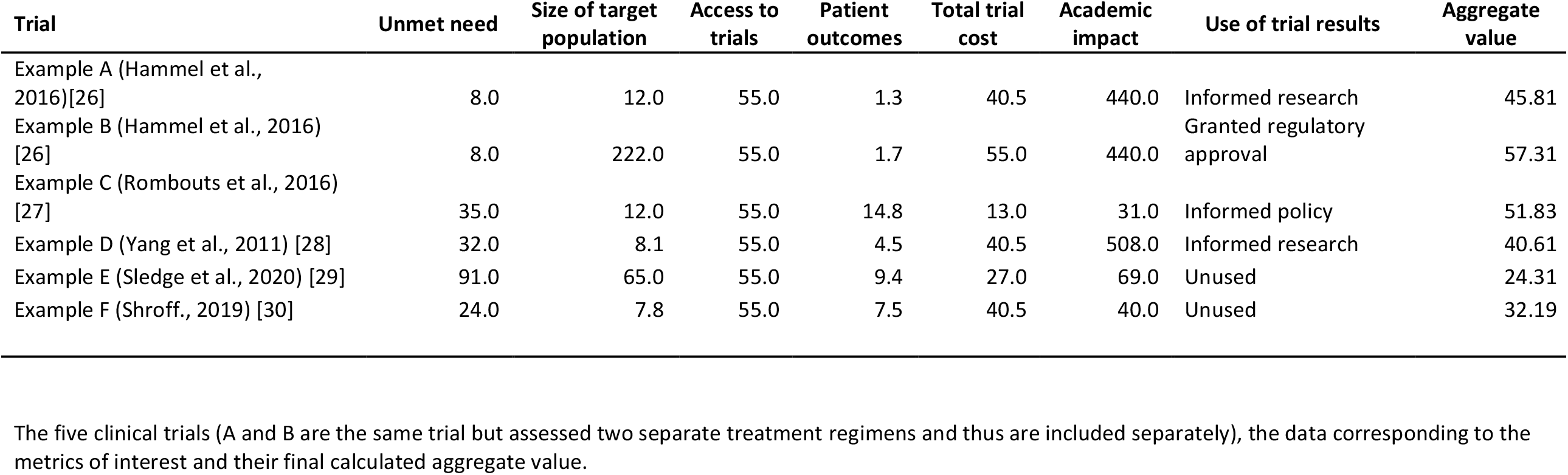
The five clinical trials (A and B are the same trial but assessed two separate treatment regimens and thus are included separately), the data corresponding to the metrics of interest and their final calculated aggregate value.

| **Trial** | **Unmet need** | **Size of target population** | **Access to trials** | **Patient outcomes** | **Total trial cost** | **Academic impact** | **Use of trial results** | **Aggregate value** |
| --- | --- | --- | --- | --- | --- | --- | --- | --- |
| Example A (Hammel et al., 2016)[26] | 8.0 | 12.0 | 55.0 | 1.3 | 40.5 | 440.0 | Informed research | 45.81 |
| Example B (Hammel et al., 2016) [26] | 8.0 | 222.0 | 55.0 | 1.7 | 55.0 | 440.0 | Granted regulatory approval | 57.31 |
| Example C (Rombouts et al., 2016) [27] | 35.0 | 12.0 | 55.0 | 14.8 | 13.0 | 31.0 | Informed policy | 51.83 |
| Example D (Yang et al., 2011) [28] | 32.0 | 8.1 | 55.0 | 4.5 | 40.5 | 508.0 | Informed research | 40.61 |
| Example E (Sledge et al., 2020) [29] | 91.0 | 65.0 | 55.0 | 9.4 | 27.0 | 69.0 | Unused | 24.31 |
| Example F (Shroff., 2019) [30] | 24.0 | 7.8 | 55.0 | 7.5 | 40.5 | 40.0 | Unused | 32.19 |

A sensitivity analysis was performed on the metric weights comparing outcomes if the combined concordant and discordant data is used, as reported above, or only the concordant data. These results are outlined in Online Resource 3.

## 3 Results

### 3.1 Final set of metrics

The following seven metrics were selected: unmet need, size of target population, eligible participants can access the trial, patient outcomes, total trial cost, academic impact and use of trial results (Table 2).

**Table 2.**
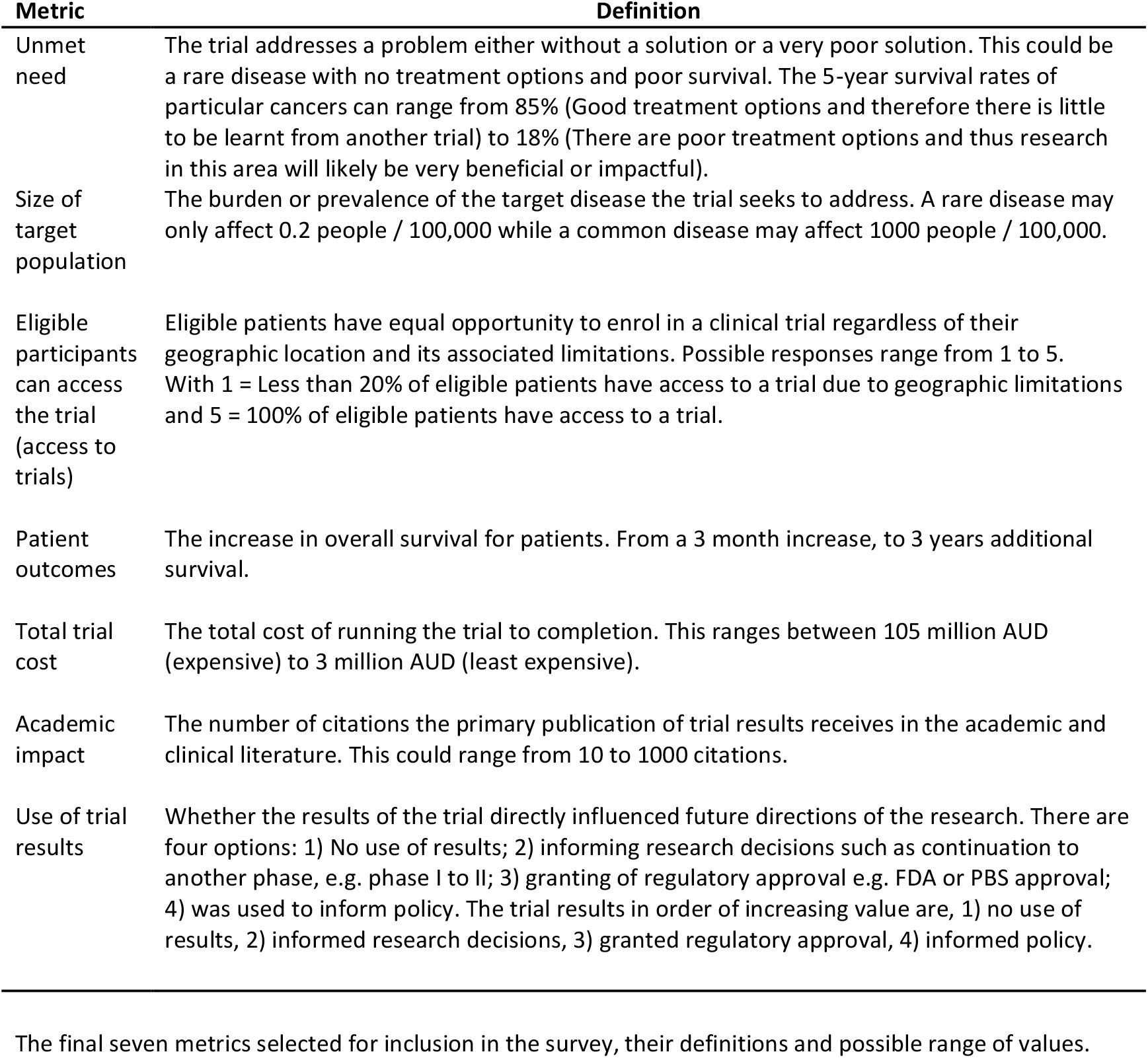
The final seven metrics selected for inclusion in the survey, their definitions and possible range of values.

### 3.2 Weight elicitation: survey responses and metric ranks

There were 157 unique consenting responses to the survey. Of those, 80 (51%) answered all the required questions. Of the participants who answered all required questions, 43 (53.8%) provided concordant responses and 37 (46.3%) provided discordant responses. Detailed characteristics of the respondents are given in **Error! Reference source not found**.3, broken down by concordant, discordant and all completed responses categories. The average standardised weights for each of the metrics using the combined data can be seen in Table 4. Unmet need had the highest average weight followed by patient outcomes. Use of trial results, size of target population and eligible participants can access the trial filled the next three positions and total trial cost and academic impact were sixth and seventh respectively. The proportion of votes each metric received for each ranking position can be seen in Figure 1. The descending order of the proportion of Rank 1 votes received by each metric is, unmet need (43.8%), patient outcomes (25%), use of trial results (13.8%), size of target population (8.8%) and access to trials (8.8%) were equal, followed by academic impact (0%) and total trial cost (0%) with no Rank 1 votes.

**Table 3.**
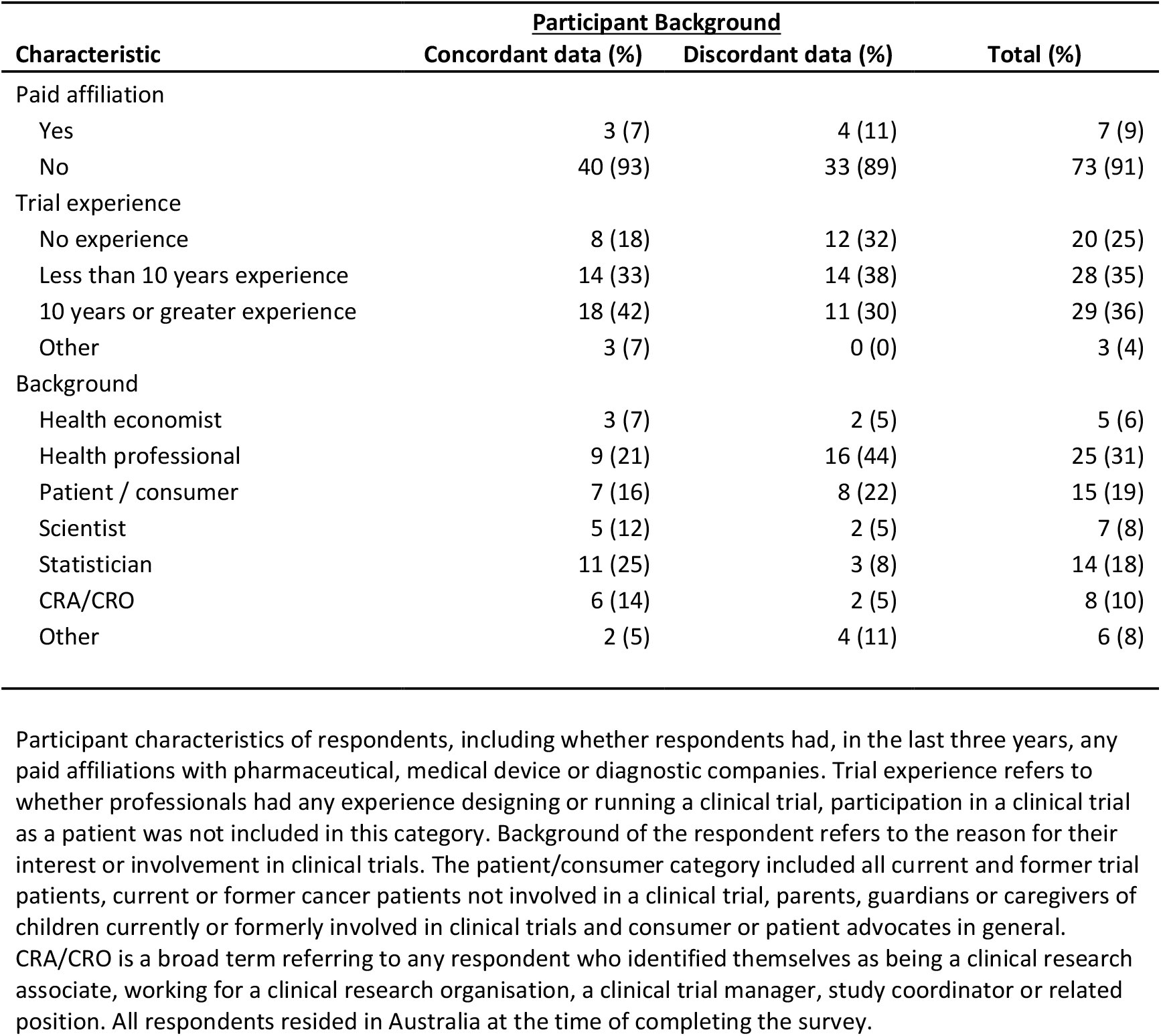
Participant characteristics of respondents, including whether respondents had, in the last three years, any paid affiliations with pharmaceutical, medical device or diagnostic companies. Trial experience refers to whether professionals had any experience designing or running a clinical trial, participation in a clinical trial as a patient was not included in this category. Background of the respondent refers to the reason for their interest or involvement in clinical trials. The patient/consumer category included all current and former trial patients, current or former cancer patients not involved in a clinical trial, parents, guardians or caregivers of children currently or formerly involved in clinical trials and consumer or patient advocates in general. CRA/CRO is a broad term referring to any respondent who identified themselves as being a clinical research associate, working for a clinical research organisation, a clinical trial manager, study coordinator or related position. All respondents resided in Australia at the time of completing the survey.

| Characteristic | Participant Background |  | Total (%) |
| --- | --- | --- | --- |
|  | Concordant data (%) | Discordant data (%) |  |
| Paid affiliation |  |  |  |
| Yes | 3 (7) | 4 (11) | 7 (9) |
| No | 40 (93) | 33 (89) | 73 (91) |
| Trial experience |  |  |  |
| No experience | 8 (18) | 12 (32) | 20 (25) |
| Less than 10 years experience | 14 (33) | 14 (38) | 28 (35) |
| 10 years or greater experience | 18 (42) | 11 (30) | 29 (36) |
| Other | 3 (7) | 0 (0) | 3 (4) |
| Background |  |  |  |
| Health economist | 3 (7) | 2 (5) | 5 (6) |
| Health professional | 9 (21) | 16 (44) | 25 (31) |
| Patient / consumer | 7 (16) | 8 (22) | 15 (19) |
| Scientist | 5 (12) | 2 (5) | 7 (8) |
| Statistician | 11 (25) | 3 (8) | 14 (18) |
| CRA/CRO | 6 (14) | 2 (5) | 8 (10) |
| Other | 2 (5) | 4 (11) | 6 (8) |

**Table 4.**
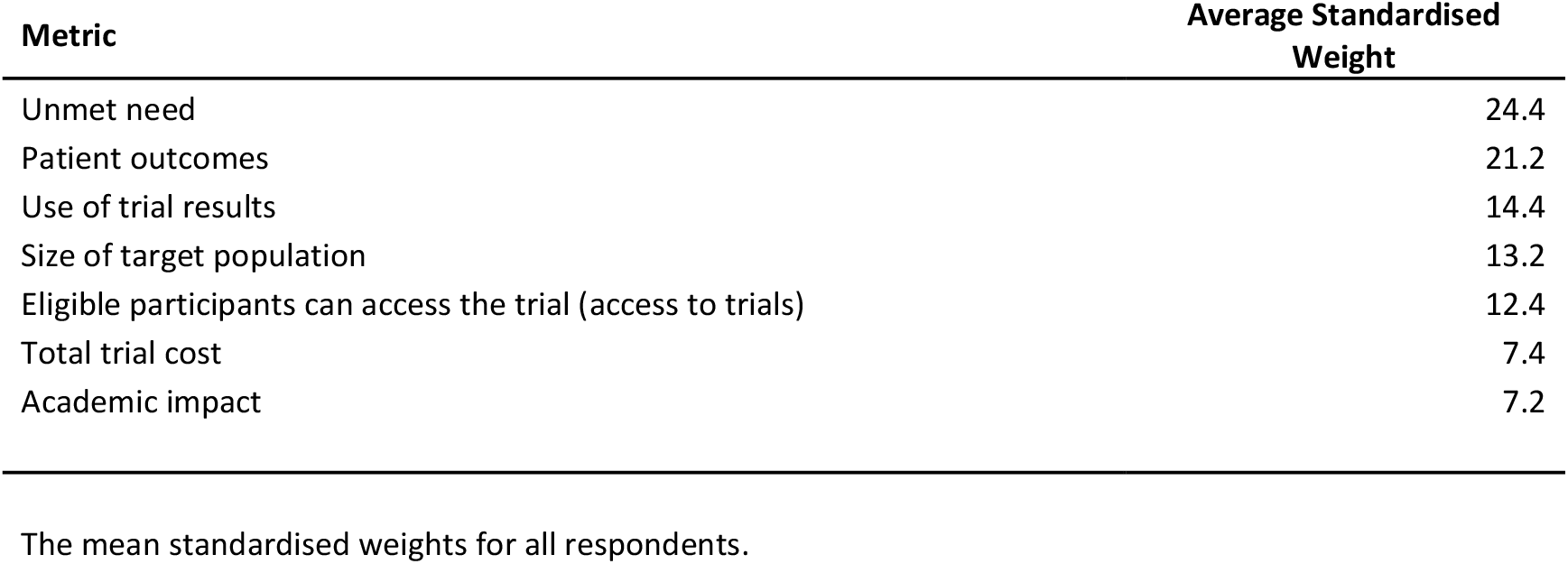
The mean standardised weights for all respondents.

**Figure 1.**
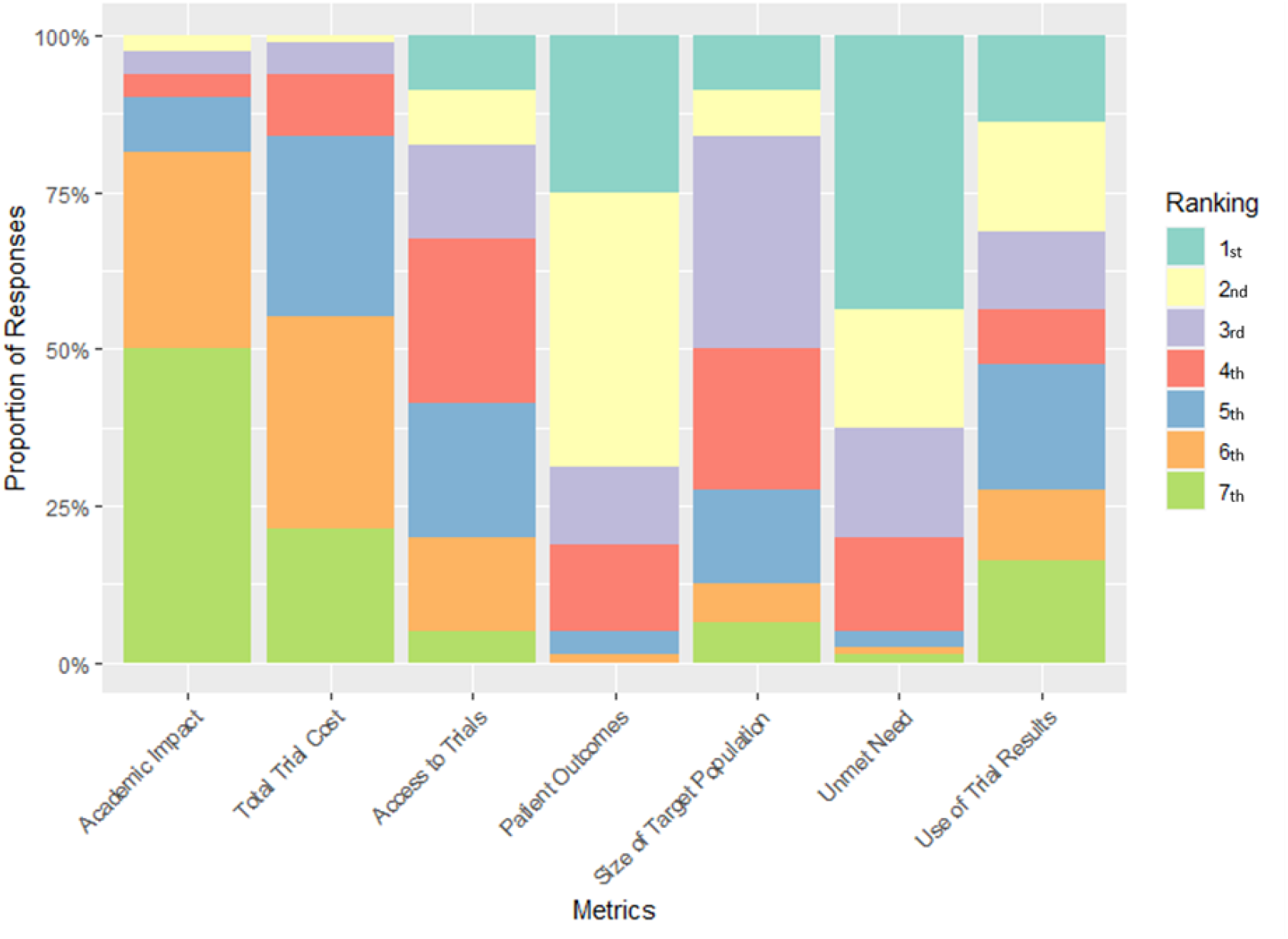
Proportion of responses each metric received for a specific ranking position

### 3.3 Application to real-world cancer clinical trials

Six cancer clinical trials were evaluated using the MCDA tool implemented via the Shiny web app. The results are displayed in Table 1. Trials were selected to cover a wide range of the included metrics. The trial with the greatest value of the example group was Hammel B 2016 with a score of 57.31, primarily due to its high ‘unmet need’. In descending order of trial value was Rombouts 2016 (51.83), Hammel A 2016 (45.81), Yang 2011 (40.61), Shroff 2019 (32.19) and Sledge 2020 (24.31).

For the six example trials, the most impactful metric driving changes in overall value is ‘unmet need’ as it contributed the most value to four of the six trials assessed.

## 4 Discussion

The results of this work demonstrate the feasibility of taking a decision analytical approach to valuing clinical trials. We believe that our MCDA decision tool is an important step toward improving the process of clinical trial prioritisation. Further, we have provided a convincing proof-of-concept through our use of real-world trial data. The same principles may be used by funders or clinical trial units to prioritise new trials, yet this requires prospective validation and reliable estimation of the trial metrics’ performance prior to trial completion. For estimated clinical outcomes, this may be a difficult process.

Due to the novelty of our work, there are no known directly comparable approaches. The closest examples and perhaps the most well-known are the ESMO magnitude of clinical benefits scale [6] the ASCO value framework [5] and using value of information for real time prioritisation decisions [31]. The ESMO framework attempts to balance the benefits of a treatment against any side effects. The ASCO framework sets out to balance treatment benefits against cost. Utilisation of value of information allowed for prospective prioritisation of phase II/III cancer clinical trials. Each respective tool has their place but, in comparison to our decision tool, each considers a clinical trial value in a highly restricted way. Additionally, each of the frameworks are focused on latter phase trials, while our tool is potentially much more broadly applicable.

Although our MCDA decision tool is developed in the context of cancer clinical trials, it could be further extended to trials covering other diseases through improvement of the ‘patient outcomes’ metric. Currently, only trials reporting a difference in overall survival between groups can be assessed. By extending the metric to enable trials that report, for example, progression-free survival, quality adjusted life years or toxicity, a more varied range of trial types could be assessed. If other clinical endpoints are to be included, such as symptom scores, it would require re-assessment of the weights for each metric.

A key strength of our method is that it explicitly incorporates the subjective preferences of a diverse group of stakeholders. It would be straightforward to adjust the metrics used to represent the values of one specific stakeholder group. Whether using preferences from diverse stakeholders or a single group, the use of our decision tool provides a transparent means to screen prospective trials at very low cost.

Our study is the first of its kind and not without limitations. A key difficulty that we encountered was the varying levels of clinical trial knowledge among participants. By presenting the survey to as many interested groups as possible we included participants who had only a limited familiarity with clinical trials. This was highlighted by the fact that a greater proportion of respondents identifying as patients/consumer advocates/family members of patients started the survey and then failed to complete it. This pattern of participant dropout may have altered the results to some extent. We believe our results are robust to this effect, but this has not been evaluated.

## 5 Conclusions

This study has demonstrated the feasibility of a broadly applicable tool for assigning value to clinical trials across a range of criteria. It is a transparent and objective tool by which to evaluate clinical trials for the purposes of prioritisation. Our hope is that the tool is used by decision makers to improve allocation of scarce medical research resources and ultimately improve patient outcomes.

## Supporting information

Online Resource 1

Online Resource 2

Online Resource 3

## Data Availability

The datasets generated and analysed during the current study are not publicly available due to consent being specific to this project with no future use permitted. Data may be made available if consent for release can be reasonably obtained from study participants.

## Acknowledgements

We would like to thank members of the steering committee Anne Woolett, Courtney Thornely, Grant McArthur, Christie Allan, Dayna Swiatek, Meredith Layton, Phil Kiossoglou, Clare Slaney and Eva Segelov for providing feedback and guidance throughout the project. We would also like to thank Anneke Grobler, Jo Cockwell, Paul Baden, Paul Fennessy, Suzanne Hasthorpe, Simon Montgomery, Sophie Broughton, Keith Donahoe and Leslie Leckie who provided feedback and their perspective on proposed metrics.

We would like to thank Atara Posner, Colin Wood, Grace Chazan, Sharon Carvalho, Michelle Tew and Sophy Athan for participating in the partial value function elicitation.

We would also like to thank all those who responded to the survey.

