## Supplementary material for "Developing and validating a multi-criteria decision analytic tool to assess the value of cancer clinical trials": Online Resource 1

Applied Health Economics and Health Policy

Piers Gillett[1], Robert K Mahar[1,2], Nancy R Tran[1], Mark Rosenthal[3,4] and Maarten IJzerman[1,3*]

Corresponding author: Maarten IJzerman, University of Melbourne Centre for Cancer Research, 305 Grattan St, Melbourne 3000 Australia.

Figure 1)

Target Pop

$y = 0.225x - 0.045 \quad [0.2, 222.67), \qquad y = 0.0643x + 35.68 \quad [222.67, 1000]$

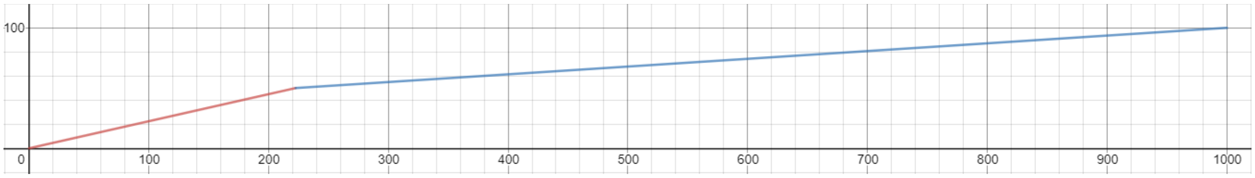

Figure 2)

Eligibility / Accessibility

$y = 1.43x - 28.6 \;\; [20, 55), \qquad y = 1.11x - 11.11 \;\; [55, 100]$

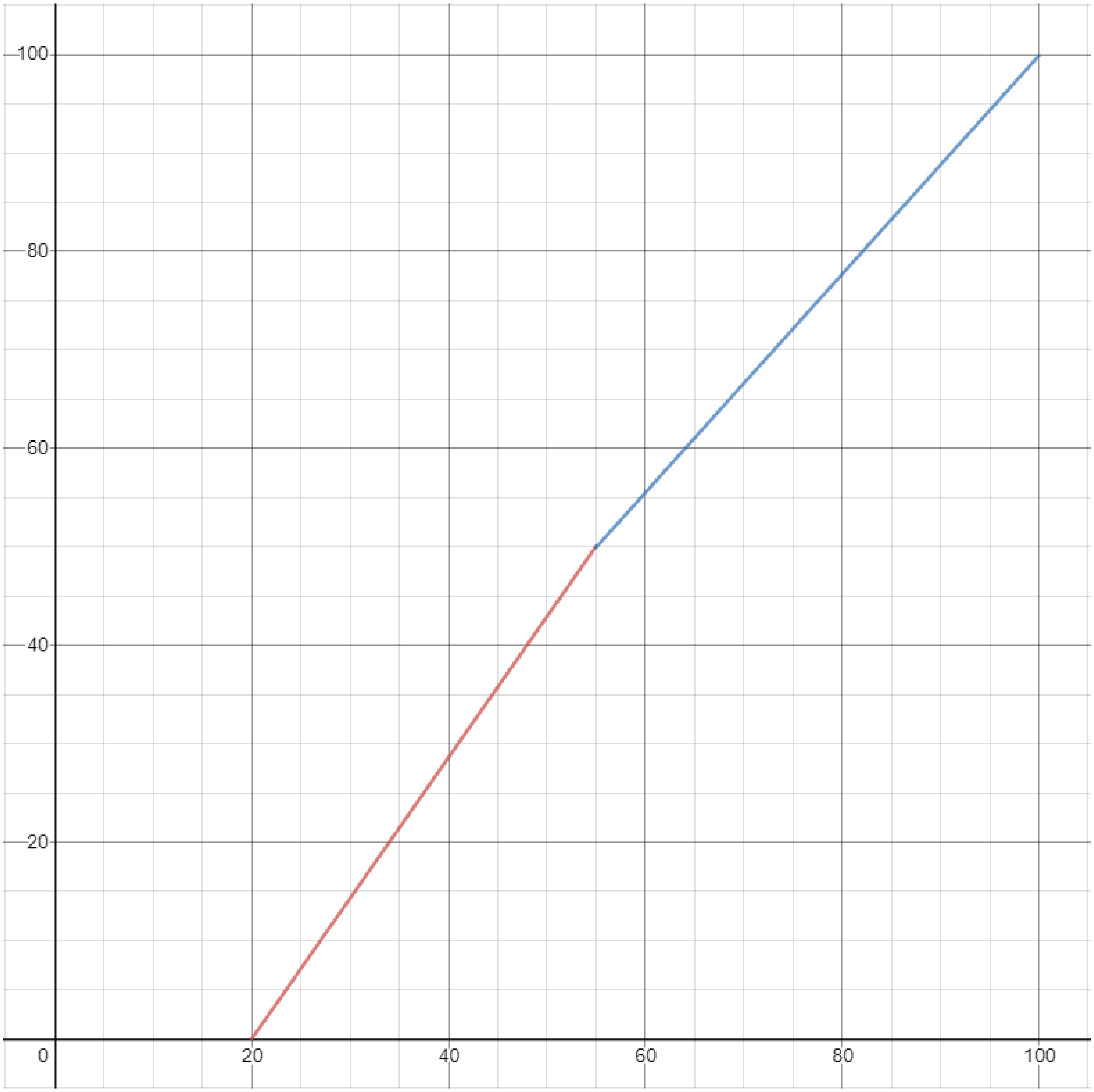

Figure 3)

Patient Outcomes

$y = 5.45x - 16.36 \;\; [3, 12.167), \qquad y = 2.10x + 24.47 \quad [12.167, 36]$

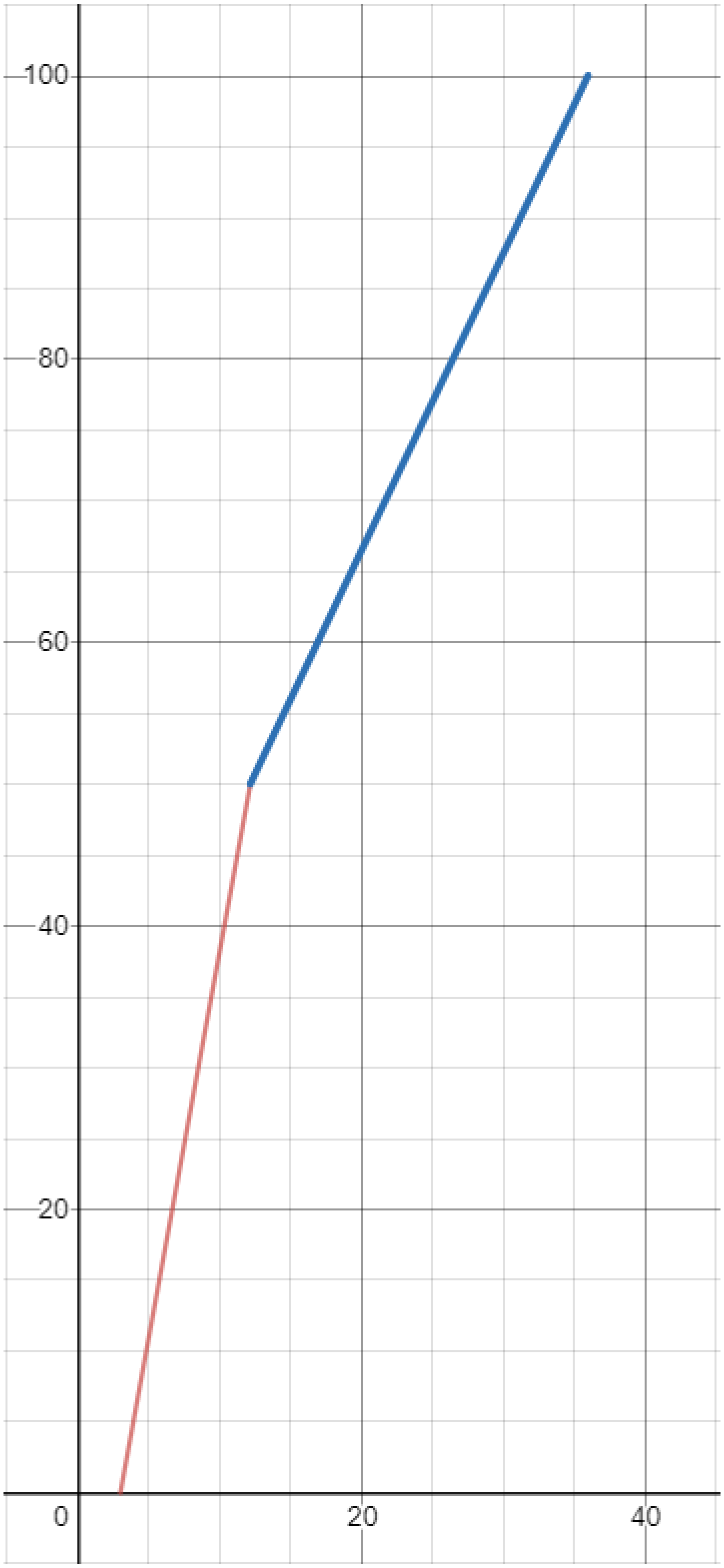

100
80
60
40
20
0
20
40

Figure 4)

Academic Impact

$$y = 0.353x - 3.53 \;\; [10, 151.67), \qquad y = 0.144x + 28.23 \;\; [151.67, 500]$$

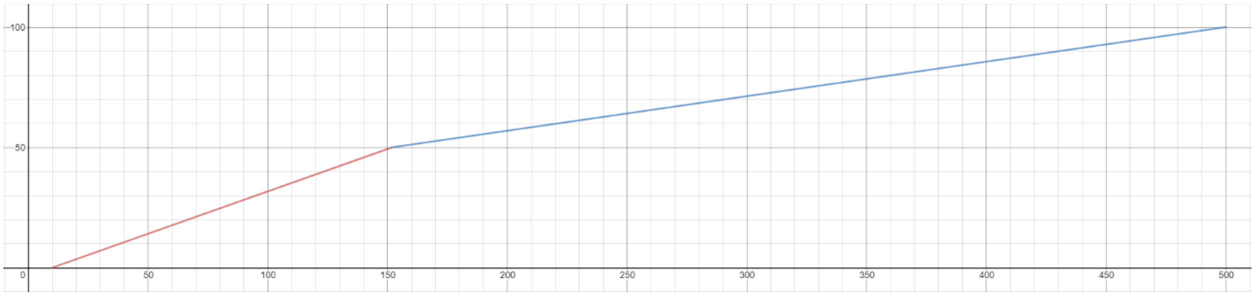

Figure 5)

Cost

$y = -0.775x + 81.4 \ \ [3, 40.5), \quad y = -1.33x + 104 \quad [40.5, 105]$

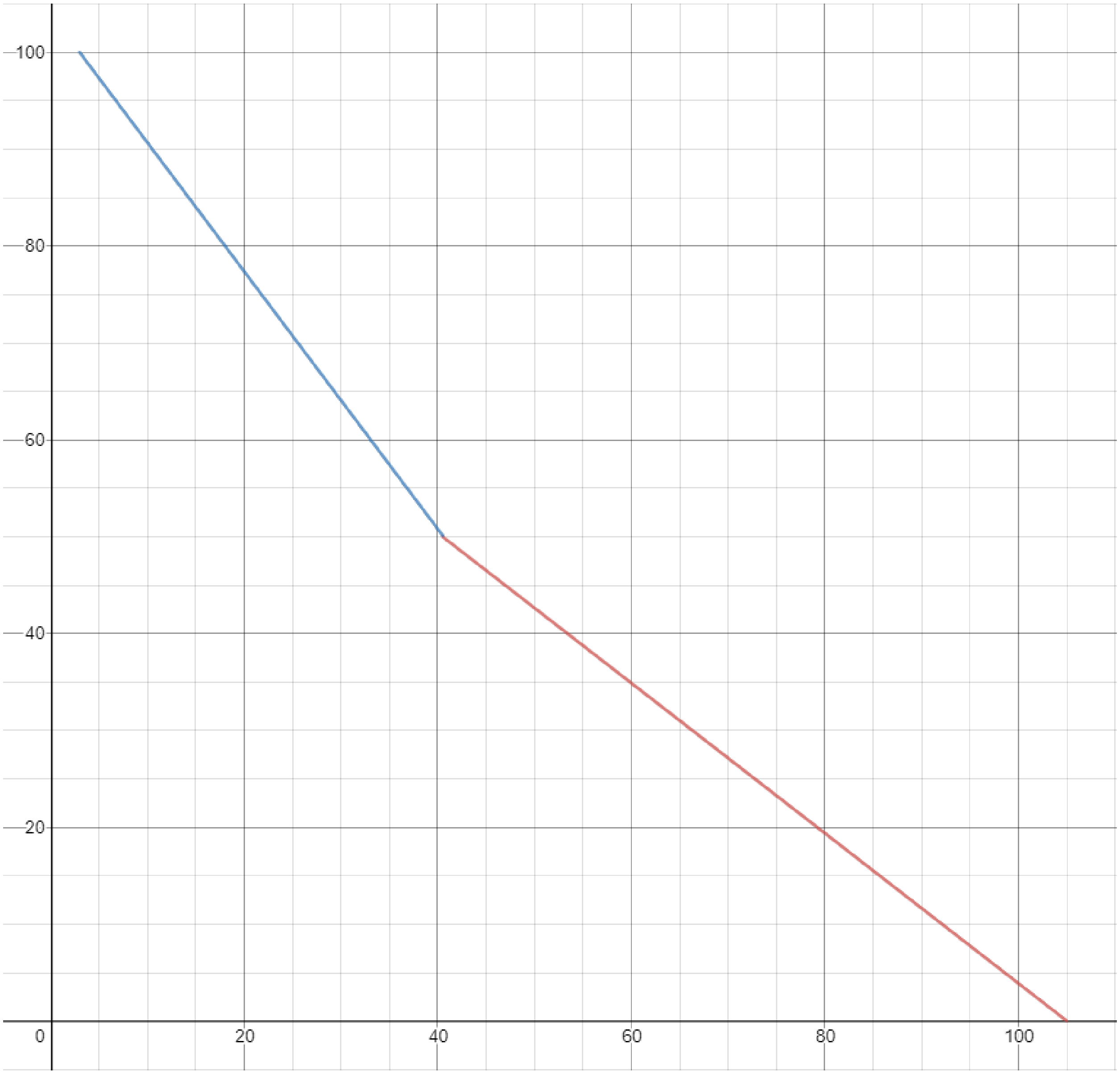

Figure 6)

Unmet Need

$y = -x + 85 \;\; [18, 35), \qquad y = -2.94x + 152.94 \qquad [35, 85]$

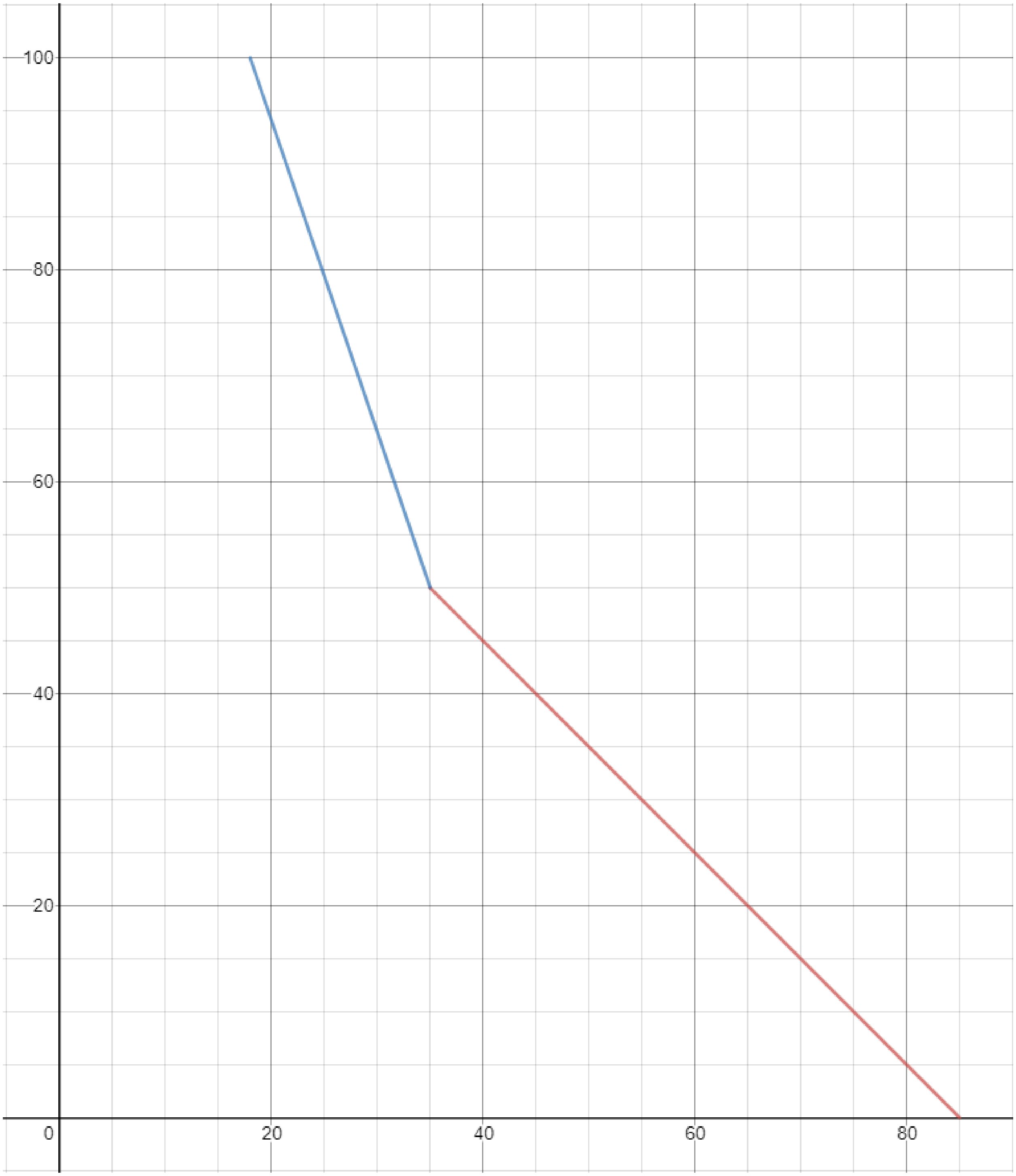
