## Supplementary material for "Developing and validating a multi-criteria decision analytic tool to assess the value of cancer clinical trials": Online Resource 2

Applied Health Economics and Health Policy

Piers Gillett[1], Robert K Mahar[1,2], Nancy R Tran[1], Mark Rosenthal[3,4] and Maarten IJzerman[1,3*]

Corresponding author: Maarten IJzerman, University of Melbourne Centre for Cancer Research, 305 Grattan St, Melbourne 3000 Australia.

Table 1)

*Table 1. The calculated total value for six example trials using the metrics weights from the concordant and combined concordant and discordant results respectively. The example trials are based upon results from published trials.*

| ***Trial*** | **Concordant Weights** | **Combined Weights** | **Weight Difference** |
|---|---|---|---|
| *Example A* | 45.8 | 48.0 | +2.2 |
| *Example B* | 57.3 | 58.6 | +1.3 |
| *Example C* | 51.8 | 52.1 | +0.3 |
| *Example D* | 40.6 | 40.2 | -0.4 |
| *Example E* | 24.3 | 21.7 | -2.6 |
| *Example F* | 32.2 | 35.8 | +3.6 |

As discussed in Step 4: Data analysis, we discovered completed responses could take one of two forms, concordant and discordant results. Steps were taken to align the data to allow for its use as outlined in the methods. A sensitivity analysis was undertaken to establish whether using the entirety of data, concordant and discordant combined, or just the concordant data would result in a difference in the calculated trial value.

Table 1 exhibits the degree of difference observed using the respective data sets. While differences were seen in the calculated trial value, the largest difference was for Example F with a difference of 3.6 points. The smallest difference was Example C with a difference of 0.3 points. Additionally, ordering these example trials from highest trial value to lowest results in the same order in both cases. Practically these differences are minor and should not change any conclusions.

In this case the sensitivity analysis identified no difference of relevance. It is conceivable, should this tool be applied that a difference in trial value of two may be enough to result in the ordering of trials changing. This tool is not intended nor designed for this level of fine detail and for all intents and purposes, the trials should be considered equal. In a case where multiple trials have very similar calculated values, if a distinction is to be made, it should be done so by relevant subject matter experts.
