## Supplementary material for "Developing and validating a multi-criteria decision analytic tool to assess the value of cancer clinical trials": Online Resource 3

Applied Health Economics and Health Policy

Piers Gillett[1], Robert K Mahar[1,2], Nancy R Tran[1], Mark Rosenthal[3,4] and Maarten IJzerman[1,3*]

Corresponding author: Maarten IJzerman, University of Melbourne Centre for Cancer Research, 305 Grattan St, Melbourne 3000 Australia.

Figure 1)

THE UNIVERSITY OF MELBOURNE

**Project:** Development and validation of a Multi-Criteria Decision Analysis based value function to determine value retrospectively of clinical trials in the Victorian Comprehensive Cancer Centre trials portfolio

Our work aims to calculate the value of a clinical trial, taking into consideration the many facets and complexities a trial may possess. This work will enable us to more accurately identify clinical trials that provide benefits to those participating in the clinical trials and provide benefit to the public at large through improved healthcare options.

The following few pages will provide you with further information about the project.

Please take the time to read this information carefully.

You may ask questions about anything you don't understand or want to know more about.

→

This project has received ethics approval from the University of Melbourne and is considered minimal risk. The full plain language statement is available below.

At the bottom of this page there is the option to provide consent to participate. You are required to give consent to participate in the survey.

---

**Plain Language Statement**
Faculty of Medicine, Dentistry and Health Sciences

**Project**: Development and validation of a Multi-Criteria Decision Analysis based value function to determine value retrospectively of clinical trials in the Victorian Comprehensive Cancer Centre trials portfolio

Thank you for your interest in participating in this research project. The following will provide you with further information about the project, so that you can decide if you would like to take part in this research.
Please take the time to read this information carefully. You may ask questions about anything you don't understand or want to know more about.
Your participation is voluntary. If you don't wish to take part, you don't have to. If you begin participating, you can also stop at any time.

The purpose of our research is to develop a decision aid. It is intended for guiding the selection of clinical trials at the Victorian Comprehensive Cancer Centre (VCCC).
We have identified a series of 'metrics' that measure different aspects of clinical trials. These metrics were identified from discussions with relevant experts and review of the academic literature. The selected metrics encompass the majority of clinical trial characteristics of relevance.

The next step in developing the decision aid is to decide the relative importance of each metric. From this survey, we will be able to calculate weightings of each metric based on your feedback.

Should you agree to participate you will complete an online survey. The survey will ask you to rank a set of metrics from 'most representative of trial value' to 'least representative'. Based on your answer in the first question, you will be asked to provide a relative weighting between the ranked metrics. The comparison weighting will allow us to determine the respective importance of each metric. The survey should take less than 15 minutes to complete.

This work will enable us to retrospectively value clinical trials. It is the first step in developing an outcome measure which we can then use in future work to try and predict the most valuable proposed trials. A tool such as this would allow for the maximisation of value of clinical trials undertaken at the VCCC. This in turn would allow for more cancer patients to access cutting edge medical care. This could mean increased survival and improved quality of life for those on the trials, as well as enhancing the medical expertise of health professionals which will improve health outcomes for all cancer patients within Victoria.

The survey does pose a minor risk of aggravating emotional trauma in individuals who have had a negative experience with a clinical trial. As the survey is entirely voluntary, you may drop out at any point without penalty while remaining anonymous.
Participation is completely voluntary. You are able to withdraw at any time. There will be no consequences for not participating. As answers are anonymous, once an answer is submitted they cannot be retracted.

Results will be published in a peer-reviewed academic journal upon completion of the project. Participants will also have the option of providing an email address which a completed manuscript will be sent to.

No personally identifiable information is required to complete the survey and thus participants will remain anonymous. Following completion of the survey, results will be stored digitally on a secure server at the University of Melbourne. Data will be kept for 5 years. Data will not be used for purposes outside of this project.

There are no conflicts of interest to report.

The project is funded by the Government of Victoria.

Ethics ID: 2056390.1
Plain Language Statement V1.0    10/03/20

If you have any questions, please feel free to contact the responsible researcher, Mr Piers Gillett.


This research project has been approved by the Human Research Ethics Committee of The University of Melbourne. If you have any concerns or complaints about the conduct of this research project, which you do not wish to discuss with the research team, you should contact the Manager, Human Research Ethics, Research Ethics and Integrity, University of Melbourne, VIC 3010. Tel: +61 3 8344 2073 or. All complaints will be treated confidentially. In any correspondence please provide the name of the research team or the name or ethics ID number of the research project.

Proceed to the next page to provide consent if you are happy to do so.

---

If you consent to participate in this survey, select the option below and click the arrow in the bottom right corner to proceed to the survey.

I consent to participate in this survey

← →

This project has received ethics approval from the University of Melbourne and is considered minimal risk. If you would like to read the full plain language statement, select the option below. If you are happy to proceed, continue to the consent page.

Show full plain language statement

Continue to consent page

←

→

**Plain Language Statement**
Faculty of Medicine, Dentistry and Health Sciences

**Project:** Development and validation of a Multi-Criteria Decision Analysis based value function to determine value retrospectively of clinical trials in the Victorian Comprehensive Cancer Centre trials portfolio

Thank you for your interest in participating in this research project. The following will provide you with further information about the project, so that you can decide if you would like to take part in this research.
Please take the time to read this information carefully. You may ask questions about anything you don't understand or want to know more about.
Your participation is voluntary. If you don't wish to take part, you don't have to. If you begin participating, you can also stop at any time.

The purpose of our research is to develop a decision aid. It is intended for guiding the selection of clinical trials at the Victorian Comprehensive Cancer Centre (VCCC).
We have identified a series of 'metrics' that measure different aspects of clinical trials. These metrics were identified from discussions with relevant experts and review of the academic literature. The selected metrics encompass the majority of clinical trial characteristics of relevance.
The next step in developing the decision aid is to decide the relative importance of each metric. From this survey, we will be able to calculate weightings for each metric based on your feedback.

Should you agree to participate you will complete an online survey. The survey will ask you to rank a set of metrics from 1 to 7 based upon the value of the swing of the metric from its worst outcome to its best outcome. You will then be asked to provide relative weights for each metric on a scale of 0 to 100. The weighting information will allow us to determine the respective importance of each metric. The survey should take less than 15 minutes to complete.

This work will enable us to retrospectively value clinical trials. It is the first step in developing an outcome measure which we can then use in future work to try and predict the most valuable proposed trials. A tool such as this would allow for the maximisation of value of clinical trials undertaken at the VCCC. This in turn would allow for more cancer patients to access cutting edge medical care. This could mean increased survival and improved quality of life for those on the trials, as well as enhancing the medical expertise of health professionals which will improve health outcomes for all cancer patients within Victoria.

The survey does pose a minor risk of aggravating emotional trauma in individuals who have had a negative experience with a clinical trial. As the survey is entirely voluntary, you may drop out at any point without penalty while remaining anonymous. As answers are anonymous, once an answer is submitted they cannot be retracted.

Results will be published in a peer-reviewed academic journal upon completion of the project. Participants will also have the option of providing an email address which a completed manuscript will be sent to.

No personally identifiable information is required to complete the survey and thus participants will remain anonymous. Following completion of the survey, results will be stored digitally on a secure server at the University of Melbourne. Data will be kept for 5 years. Data will not be used for purposes outside of this project.

There are no conflicts of interest to report.

The project is funded by the Victorian Comprehensive Cancer Centre.

Ethics ID: 2056390.1
Plain Language Statement V2.0     21/04/20

If you have any questions, please feel free to contact the responsible researcher, Mr Piers Gillett.


This research project has been approved by the Human Research Ethics Committee of The University of Melbourne. If you have any concerns or complaints about the conduct of this research project, which you do not wish to discuss with the research team, you should contact the Manager, Human Research Ethics, Research Ethics and Integrity, University of Melbourne, VIC 3010. Tel: +61 3 8344 2073 or. All complaints will be treated confidentially. In any correspondence please provide the name of the research team or the name or ethics ID number of the research project.

Proceed to the next page to provide consent if you are happy to do so.

←  →

THE UNIVERSITY OF
MELBOURNE

If you consent to participate in this survey, select the option below and click the arrow in the bottom right corner to proceed to the survey. You can only proceed if you provide consent.

I consent to participate in this survey

←  →

THE UNIVERSITY OF MELBOURNE

What is your involvement in clinical trials?

Please select the option that best describes the **majority** of your involvement with clinical trials if multiple options are applicable.

- Patient
- Health Professional
- Health Regulator
- Health Economist
- Scientist (Basic Research)
- Statistician
- Other

←  →

THE UNIVERSITY OF MELBOURNE

What is your involvement with clinical trials?

THE UNIVERSITY OF MELBOURNE

Do you currently have or have you held, within the last 3 years, any paid affiliations with any pharmaceutical, medical device or diagnostics companies?

- Yes
- No

THE UNIVERSITY OF MELBOURNE

Do you have any experience designing and/or running a clinical trial?

- No, I have no experience designing or running a clinical trial
- Yes, I have some experience but less than 10 years experience designing and/or running a clinical trial.
- Yes, I have more than 10 years experience designing and/or running a clinical trial.
- Other

THE UNIVERSITY OF MELBOURNE

Please describe your level of experience with clinical trials.

THE UNIVERSITY OF
MELBOURNE

In which country do you currently live?

THE UNIVERSITY OF
MELBOURNE

The questions on the next page will ask you about the metrics (measurable characteristics of trial) that you believe are most important in representing the value of a clinical trial.

The definitions of these metrics and their possible range of values are as follows:

**Unmet Need** - The trial addresses a problem either without a solution or a very poor solution. This could be a rare disease with no treatment options and poor survival. The 5-year survival rates of particular cancers can range from 85% (Good treatment options and therefore there is little to be learnt from another trial) to 18% (There are poor treatment options and thus research in this area will likely be very beneficial or impactful).

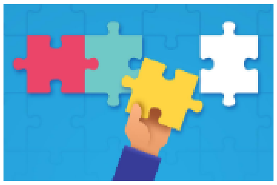

**Size of Target Population** - The burden or prevalence of the target disease the trial seeks to address. A rare disease may only affect 0.2 people / 100,000 while a common disease may affect 1000 people / 100,000.

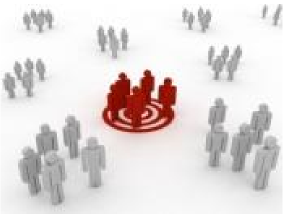

**Eligible participants can access the trial** - Eligible patients have equal opportunity to enrol in a clinical trial regardless of their geographic location and its associated limitations. Possible responses range from 1 to 5. With 1 = Less than 20% of eligible patients have access to a trial due to geographic limitations and 5 = 100% of eligible patients have access to a trial.

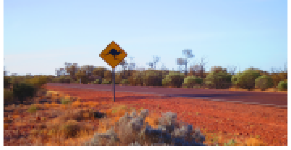

**Patient Outcomes** - The increase in overall survival for patients. From a 3 month increase, to 3 years additional survival.

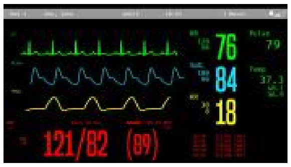

**Total Trial Cost** - The total cost of running the trial to completion. This ranges between 105 million AUD (expensive) to 3 million AUD (least expensive).

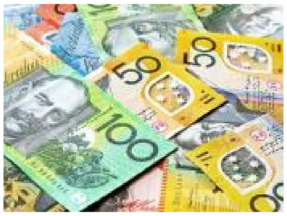


**Academic Impact** - The number of citations the primary publication of trial results receives in the academic and clinical literature. This could range from 10 to 1000 citations.

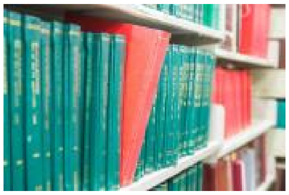

**Use of Trial Results** - Whether the results of the trial directly influenced future directions of the research. There are four options: 1) No use of results; 2) informing research decisions such as continuation to another phase, e.g. phase I to II; 3) granting of regulatory approval e.g. FDA or PBS approval; 4) was used to inform policy. The trial results in order of increasing value are, 1) no use of results, 2) informed research decisions, 3) granted regulatory approval, 4) informed policy.

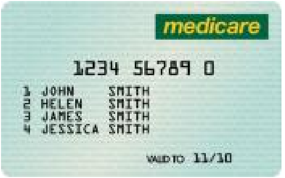


Due to its paramount importance, it is assumed a trial is appropriately designed so as to ensure statistical validity and confidence in the results. Therefore, there is no quality of trial design metric included as an option.

THE UNIVERSITY OF MELBOURNE

This question requires you to rank a list of metrics that represent different components of clinical trials. Simply drag and drop the metrics to place them in your desired order.

Using the range of values provided for each metric, please rank the metrics from most to least important according to the value of each, if the metric was to change from its worst outcome to its best outcome. For example, compare the swing from 10 citations to 500 citations of the primary trial publication to a swing of 0.2 people / 100,000 to 1000 people / 100,000 for the size of target population. The swing or change you consider to add more value should be ranked higher.

The worst and best outcomes of each metric are provided with metrics below.

**Unmet Need** - The 5-year survival rates of particular cancers can range from 85% to 18%.

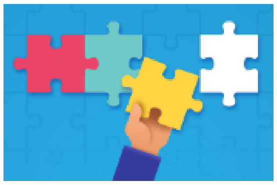

**Size of Target Population** - A rare disease may only affect 0.2 people / 100,000 while a common disease may affect 1000 people / 100,000.

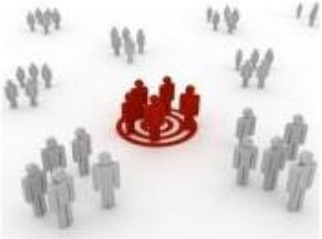

**Eligible participants can access the trial** - Possible responses range from 1 to 5. With 1 = Less than 20% of eligible patients have access to a trial due to geographic limitations and 5 = 100% of eligible patients have access to a trial.

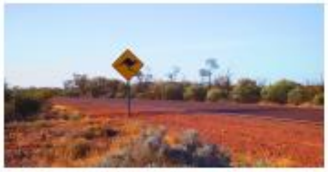

**Patient Outcomes** - From a 3 month increase, to 3 year increase in survival.

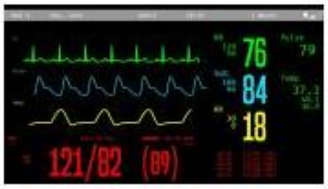


**Total Trial Cost** - This ranges between 105 million AUD to 3 million AUD.

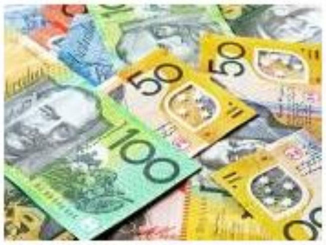

**Academic Impact** - This could range from 10 to 1000 citations.

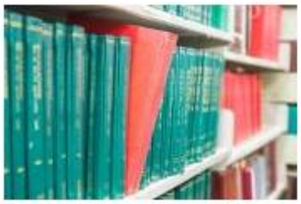

**Use of Trial Results** - The trial results in order of increasing value are, 1) no use of results, 2) informed research decisions, 3) granted regulatory approval, 4) informed policy.

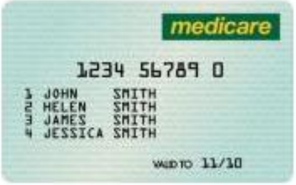

In addition to the rankings of the metrics, we would like to capture your opinions on the relative importance or weights of the metrics.

For the metric you ranked first, please assign it a value of 100. For all following metrics, provide a value between 1 and 99 with the lower the ranking, the lower the value. For example, the top ranked metric receives a score of 100. If you believe the second ranked metric is 20% less valuable, give it a score of 80. Continue this process until you have provided weights for all the metrics.

**Please note that the order of metrics in this list is not updated based upon your previous rankings. Be careful to make sure the correct metric receives your intended weighting.**

0 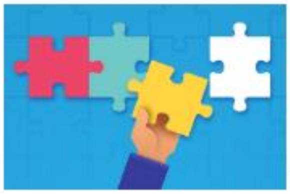 **Unmet Need**

0 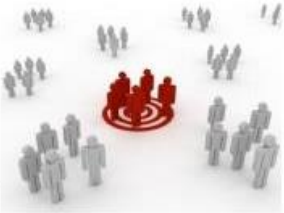 **Size of Target Population**

0 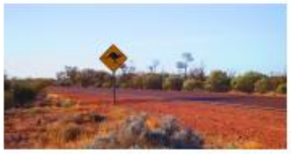 **Eligible participants can access the trial**

0 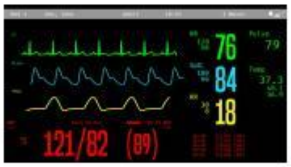

**Patient Outcomes**

0 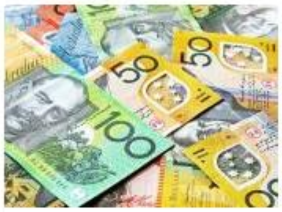 Total Trial Cost

0 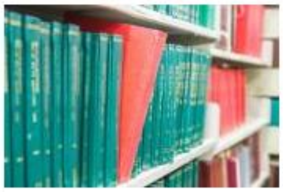 Academic Impact

0 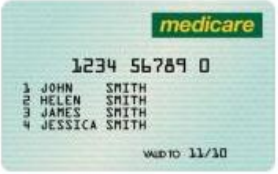

Use of Trial Results

THE UNIVERSITY OF
MELBOURNE

If you have any additional feedback about the survey, the metrics, valuation of clinical trials or anything else you think may be helpful please provide it here.

(Optional)

If you would be interested in receiving the final report, please leave a contact email and your name below.

(Optional)

We thank you for your time spent taking this survey.
Your response has been recorded.
